# Development and Feasibility of a Communication Accommodation Training Module for Health Profession Students

**DOI:** 10.64898/2026.10.16.26365768

**Authors:** Kathrin Rothermich, Rebecca Clark-Stallkamp, Rose P. Baker-Iyore, Charlene Cuatico, Joseph G. L. Lee, Susan C. Bobb

**Affiliations:** Department of Communication Sciences and Disorders, East Carolina University, Greenville, NC, USA; Department of Interdisciplinary Professions, College of Education, East Carolina University, Greenville, NC, USA; Independent instructional design consultant, Reading, MA, USA; Department of Implementation Science, Division of Public Health Sciences, Wake Forest University School of Medicine, Winston-Salem, NC, USA; Department of Psychology, Gordon College, Wenham, MA, USA

**Keywords:** Communication Accommodation, Health Professions Education, Cultural Responsiveness, Communication Training, Speech-Language Pathology

## Abstract

**Objectives:** Health professions students often receive limited training on how to adjust their communication for patients whose first language is not English (LX), despite the risks for diagnosis, treatment, and patient trust. Based on Communication Accommodation Theory, we developed a brief module to improve students’ awareness and knowledge of strategic communication adjustments for LX patients.

**Methods:** We tested feasibility in a single-group, pre-post pilot study guided by the Kirkpatrick framework with undergraduate (n=22) and graduate (n=41) speech-language pathology and psychology students in Massachusetts and North Carolina. The module included instructional content, video and audio examples, interactive activities, and a podcast. Learning was assessed with knowledge checks, and self-reported facilitative and resistant accommodation with the Communication Accommodation Behavior Scale (CABS). After training, students rated their confidence and likelihood of using accommodation strategies.

**Results:** At both sites, facilitative accommodation increased and resistant accommodation decreased significantly, with a larger decrease at the undergraduate site. Knowledge-check accuracy was high at both time points, suggesting a ceiling effect. Students reported moderate-to-high confidence in adapting their speech and high likelihood of using a learned strategy in future clinical encounters.

**Conclusions:** A brief module can be feasibly delivered across institutions and is associated with short-term improvements in self-reported communication accommodation.

**Practice implications:** We recommend future work evaluate whether these changes translate into clinical behavior long-term and refine knowledge-check items to better capture learning gains.

## 1. Introduction

In the US, roughly 1 in 5 people speak a language other than English at home [1] and healthcare providers are seeing an increased number of patients whose first language is not English. While access to medical interpreters is required under federal law [2], implementation is not consistent, and some patients may prefer not to use an interpreter. When a patient and their provider prefer different languages, it can become harder to have conversations about symptoms and follow-up care [3,4] because they are making decisions under uncertainty [5]. For example, patients whose first language is not the dominant language (hereafter, “LX” speakers) may struggle to communicate what hurts, and providers may struggle to explain what symptoms mean [6,7]. These communication problems may lead to misdiagnosis, medical errors, mistrust, and poor health outcomes [8]. When a patient cannot fully express themselves, or sense their provider is impatient, they might also be less likely to share important information [3,9,10]. Recent work often focuses on provider interactions with interpreters and less on direct patient care with LX speakers (e.g., [11]). Health professions students also report receiving little formal instruction about how to communicate with LX patients [12]. Here, we focus on how we could train providers to more effectively communicate with LX patients.

One way to improve patient-provider communication is through communication accommodation. Communication Accommodation Theory (CAT) explains how speakers adjust their speech, tone, and gestures to align with a conversation partner [13]. Two strategies drive this process: convergence, where speakers reduce linguistic differences to build rapport, and divergence, where they maintain or increase those differences [14]. In healthcare interactions, accommodation often takes the form of acoustic speech adjustments (speech accommodation), such as slowing speech rate, hyperarticulation, or modified pitch, and lexical adjustments, such as using simpler vocabulary and sentence structure [15]. Adequate accommodation is generally seen as helpful, but speakers can also misjudge the level needed based on appearance, language production proficiency, and biases. These judgments are often made under conditions of uncertainty [16], as speakers typically have incomplete information about a listener’s communication needs, comprehension, or preferences. Therefore, a speaker might change their speech more than necessary (overaccommodation) which can come across as condescending. Underaccommodation happens when a speaker changes too little, leaving the listener without support [17]; both patterns are linked to strained communication and avoidance [18].

One approach to addressing these communication challenges is to improve the preparation of future clinicians regarding patient-provider communications [19]. While cultural responsiveness and competency are now standard components of many health profession curricula [20], they often leave out how to best communicate with LX patients. Although some programs incorporate interpreter training or simulation experiences [21], relatively few focus on how clinicians can directly adapt their communication when interacting with LX patients [11,22]. Online educational modules may offer a scalable approach to addressing this gap. Here, we describe the development and feasibility of a training module that includes video and audio materials, group discussions, and self-assessment activities. The module also incorporates stimuli adapted from our avatar-based simulation study with physician assistant students [15]. To assess feasibility across different student populations, we piloted the module in two courses, one undergraduate and one graduate, at two universities. We aimed to (1) assess the feasibility of delivering the module across two institutions in terms of uptake and completion and (2) explore preliminary changes in learner reactions, knowledge, and self-reported accommodation intentions, guided by the Kirkpatrick model [23].

## 2. Methods

### 2.1. Intervention reporting

Reporting follows the Criteria for Reporting on Development and Evaluation of Professional Training interventions in Healthcare (CRe-DEPTH) [24]; see Supplementary File 1.

### 2.2. Participants

This study had two recruiting sites. We recruited students (*n* = 24) as part of an undergraduate speech-language pathology or psychology program at Gordon College, as the course enrolls students from both majors who are preparing for clinical or health-related careers. Students were in the second, third, or fourth year of their program. We also recruited graduate students (*n* = 45) in a speech-language pathology master’s program as part of a second-year course at East Carolina University. The East Carolina University Institutional Review Board approved this study under an exempt determination (UMCIRB 25-001427). Participating students provided written informed consent to take part in the study. Students completed the activity as part of their regular course requirements and did not receive extra credit. Students at both locations were given two options: 1) complete the *Communication in Healthcare: Limited English Proficiency Contexts* training module or 2) complete an alternative assignment consisting of two videos on limited English proficiency and federal guidelines, followed by a reflection on their relevance to the student’s life or future career. Both options fulfilled the same course requirement. One student at East Carolina University opted out of the research portion and was provided with the alternative assignment.

Pre- and post-module responses were matched using participant-generated anonymous identifiers. Five participants were excluded from analysis: three completed only the pre-module survey (two at East Carolina University, one at Gordon College), one Gordon College participant completed only the post-module survey, and one East Carolina University participant completed the post-module survey before the pre-module survey, so their responses could not reflect completion of the module. The final analytic sample was 63 (Gordon College *n* = 22; East Carolina University *n* = 41).

Table 1 shows demographic characteristics of the final sample. Most participants were female, and ages ranged from 19 to 52.

**Table 1.**
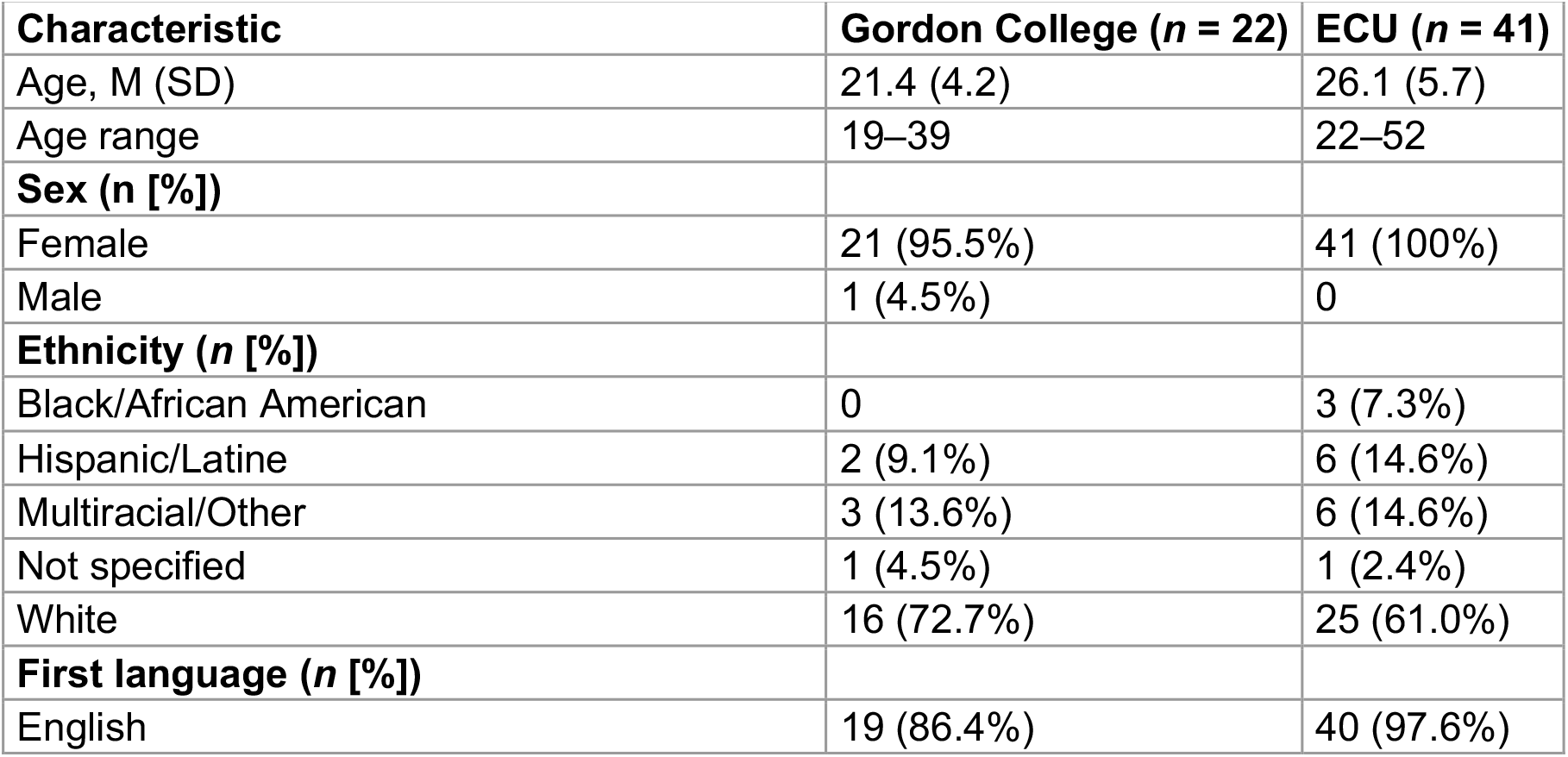
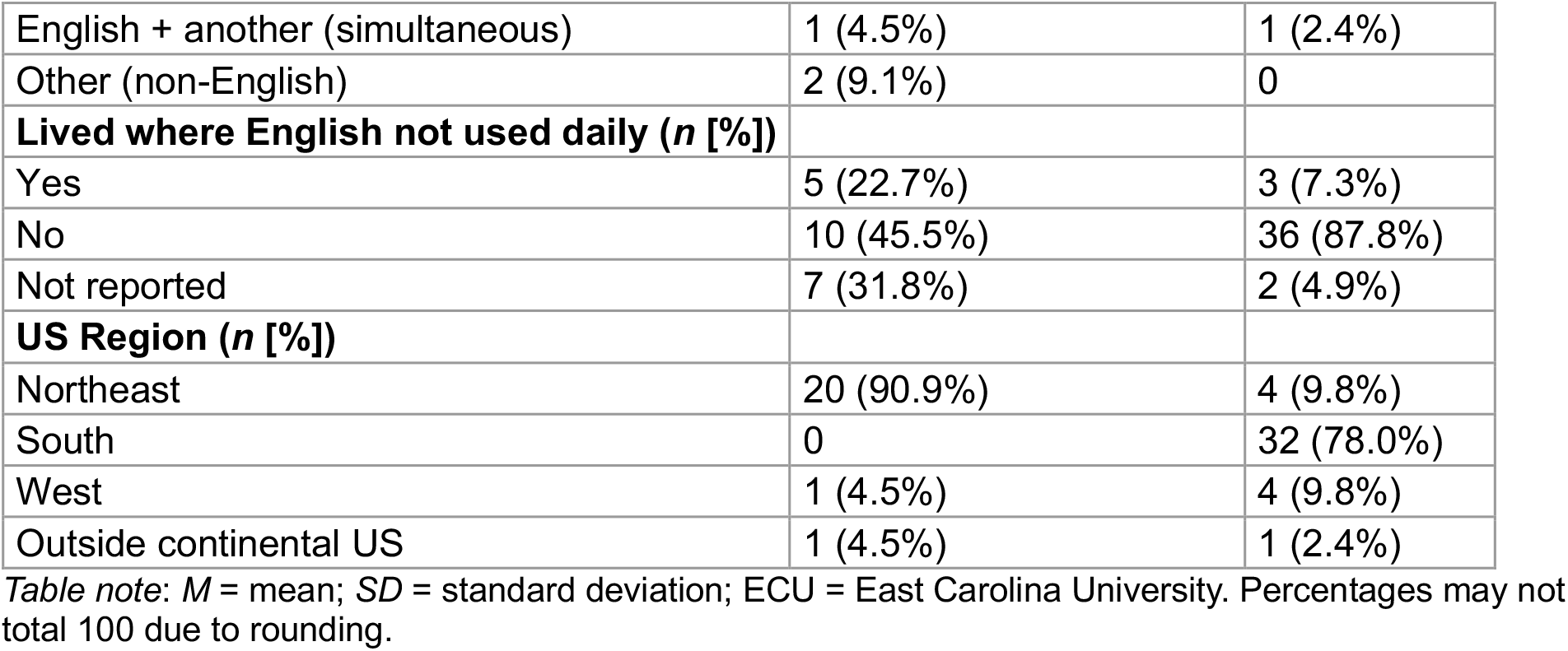
Characteristics of participants in the module evaluation in fall of 2025 (n=63).

### 2.3. Module description

The online training module, *Communication in Healthcare: Limited English Proficiency Contexts*, was delivered through the Canvas Learning Management System (LMS). It introduced communication accommodation as the way speakers adjust their speech when interacting with individuals who have lower proficiency in a shared language, with the aim of helping future clinicians recognize, understand, and apply these adjustments in healthcare interactions. The module was a single, asynchronous, self-paced session of approximately 30 minutes with no follow-up, completed within one-week window in October 2025. It combined instructional content, video and audio examples, interactive activities, guided reflection, and assessment to build knowledge and skills through education, modeling, and applied practice. Content was identical across sites and academic levels. Course instructors introduced the module and posted reminders but did not give feedback on student responses. Retrospectively, these components map onto the capability component of the COM-B framework [25]; the module was not developed using a formal behavior change framework.

### 2.4. Intervention (Module) Development

Module development occurred in four iterative stages. First, an interdisciplinary team with expertise in instructional design, implementation science, communication sciences and disorders, and psychology defined the target learners, instructional goals, and scope, drawing on Communication Accommodation Theory, prior accommodation research, and existing educational resources. Second, the team worked with an instructional design consultant to storyboard and sequence the content and to develop reflective and practice activities, supported by graphics, audio, video, and interactive elements. Some video examples were adapted from an earlier avatar-based simulation study with physician assistant students [15]. Third, the module was built in Canvas and revised through iterative review of content accuracy, instructional coherence, usability, and technical functionality. Finally, evaluation was integrated throughout development by aligning instructional content with pre- and post-training measures of learning and self-reported accommodation behavior.

### 2.5. Module structure, content, and assessment

The module was developed using an ADDIE-informed (Analyze, Design, Develop, Implement, Evaluate) process, with the Kirkpatrick Evaluation Model incorporated prospectively to align instructional activities with intended learning and behavioral outcomes. As shown in **Figure 1**, evaluation considerations were integrated during **A**nalysis and **D**esign, carried through **D**evelopment and **I**mplementation, and used to structure the final **E**valuation of the intervention. The present study examined Kirkpatrick Level 1 (reaction), Level 2 (learning), and Level 3 (self-reported behavior) outcomes. Level 1 was approximated by perceived confidence and intended use, and Level 3 by self-reported anticipated behavior. Feasibility was assessed as reach of each module component and completion.

**Figure 1.**
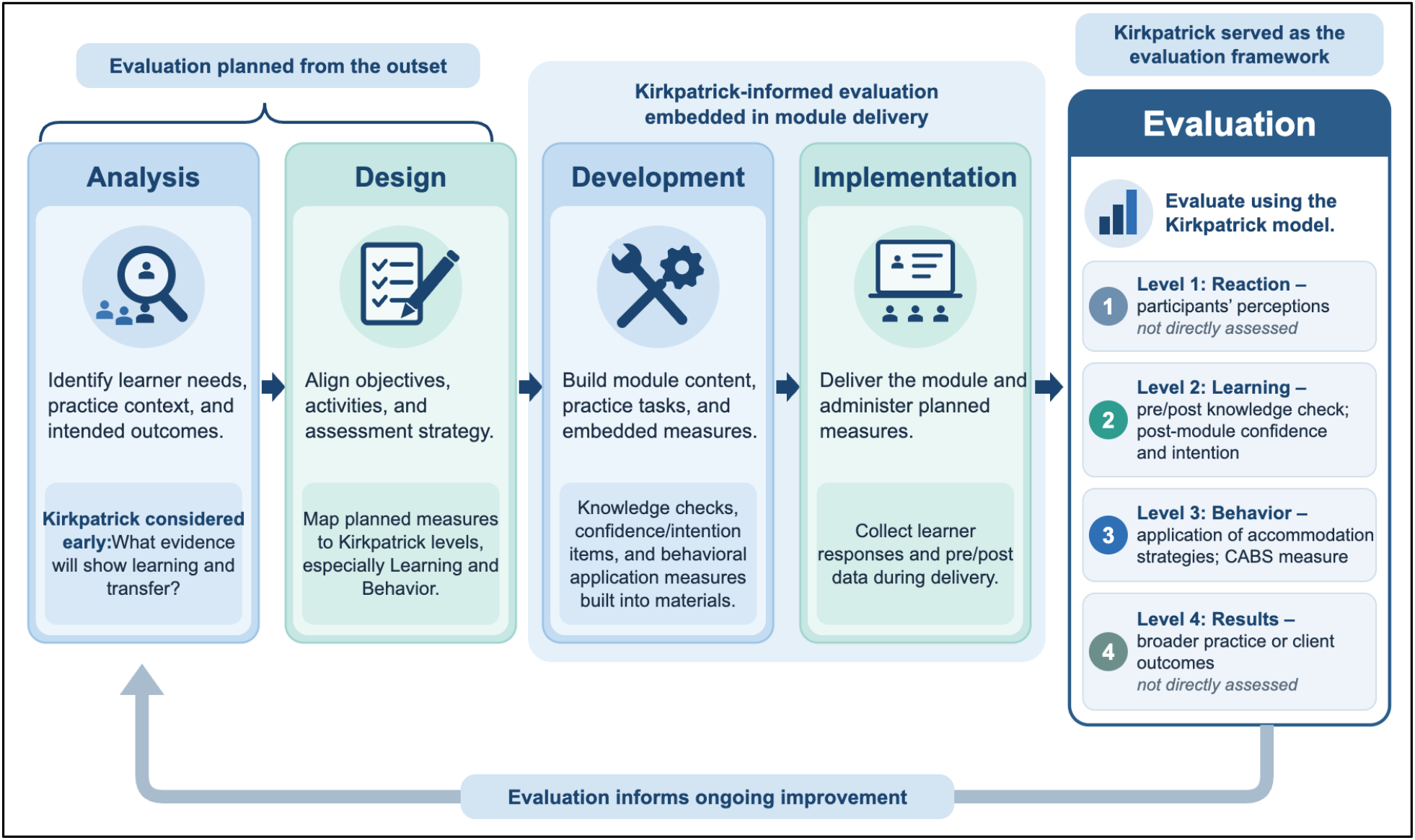
Integrating the Kirkpatrick Model into ADDIE-Based Module Development. This graphic was created using author prompts and ChatGPT (5.6 SOL). ADDIE = Analyze, Design, Develop, Implement, Evaluate, CABS = Communication Accommodation Behavior Scale.

#### 2.5.1. Structure and Procedure

The module was organized into four primary components: an introduction (1), a pre-assessment (1.1), the training content (2 & 3), and a post-assessment (4; See Canvas LMS layout in **Figure 2**). The introduction provided an overview of the training and presented four learning outcomes focused on describing typical characteristics of communication accommodation, explaining its function when differences in language proficiency exist, recognizing its use and relevance in healthcare interactions, and applying effective accommodation strategies.

**Figure 2.**
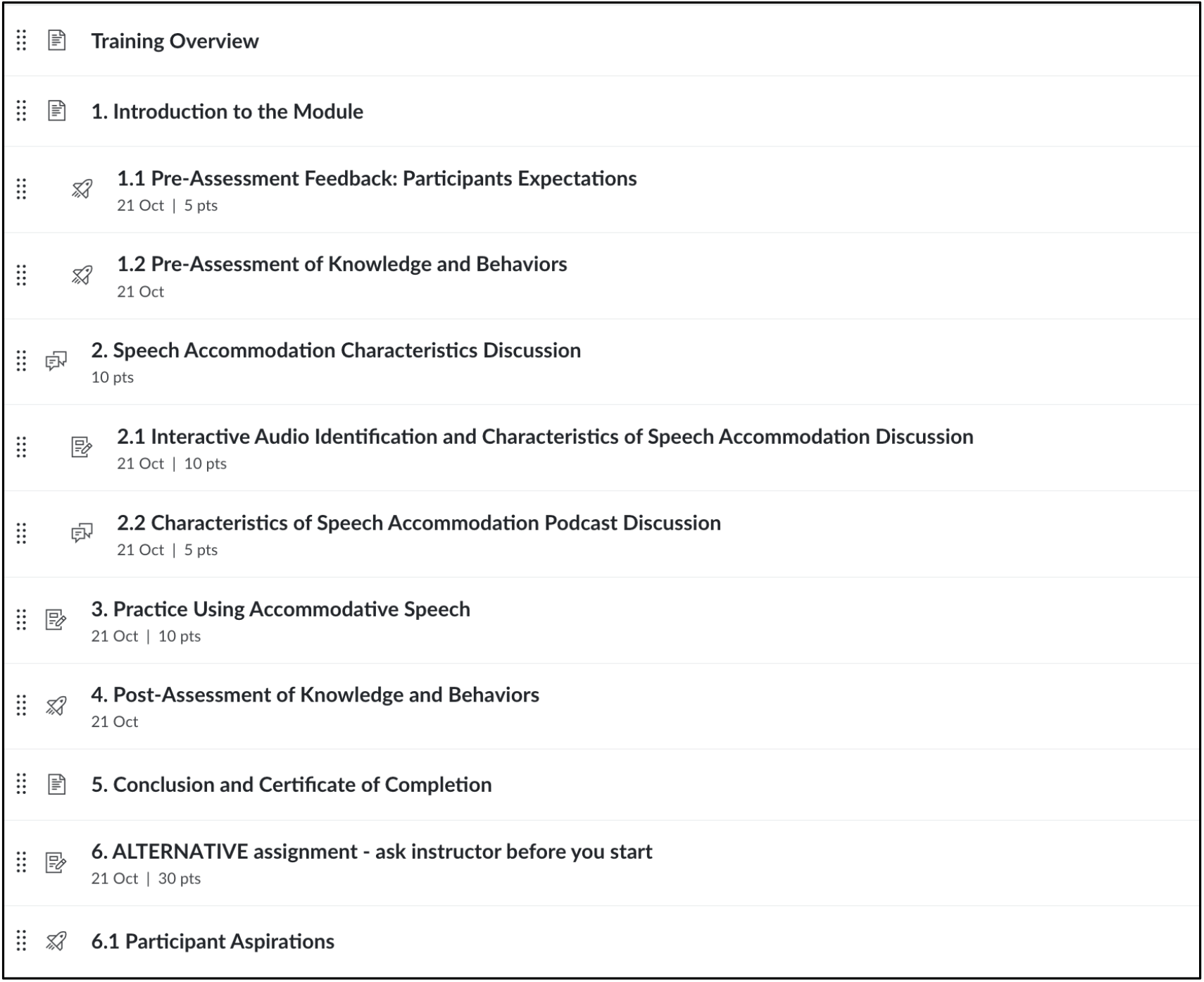
Module Structure in Canvas Learning Management System.

Before beginning the instructional content, students created a self-generated anonymous identification code to ensure pre- and post-measures could be tracked by participant. Students also completed the 8-item Communication Accommodation Behavior Scale as pre-post measure (CABS) [26], and provided demographic information related to age, sex, ethnicity, language background, time spent in non-English-speaking settings, self-rated language proficiency (based on the Language and Social Background Questionnaire [27]), and U.S. region. Students also completed four multiple-choice knowledge questions (see measurement section below for details) assessing common communication accommodation concepts and strategies.

#### 2.5.2. Content

The training content contained multiple forms of media and guided reflection to move students from observation to identification and application of accommodation strategies. Specifically, students first viewed a healthcare interaction involving a patient, nurse, physician, and interpreter and responded to prompts about how the healthcare providers adjusted their communication, which behaviors appeared helpful, and how differences in language proficiency influenced the interaction. Students then listened to four brief speech samples and identified each as accommodative or nonaccommodative. Feedback prompted the student to consider specific speech characteristics that informed their judgments, including speech rate, emphasis, and word choice. This was followed by an approximately 7-minute podcast that the authors developed using AI-assisted NotebookLM based on selected research on speech accommodation. The podcast was followed by a Canvas discussion activity designed to encourage reflection and application of the material. Students were asked to respond to one of three prompts addressing 1) what they learned from the podcast, 2) whether the examples would influence how they communicate with patients who use English as a second language, and 3) how they would determine whether a speech sample represented appropriate accommodation. A final statement encouraged students to consider accommodation as a continuum rather than a simple accommodative/non-accommodative distinction.

The final training activity provided an opportunity for students to practice accommodative speech. Using patients featured in the examples as a basis for the activity, students generated and recorded follow-up questions that they might use in a clinical interaction (**Figures 3** and **4**). They then listened to their own recordings and reflected on how their communication might be perceived by listeners with different levels of English proficiency. Reflection prompts asked students to evaluate how they sounded, consider whether their speech might be experienced differently by a less proficient English speaker, and identify ways they could modify their communication to improve both comprehensibility and rapport.

**Figure 3.**
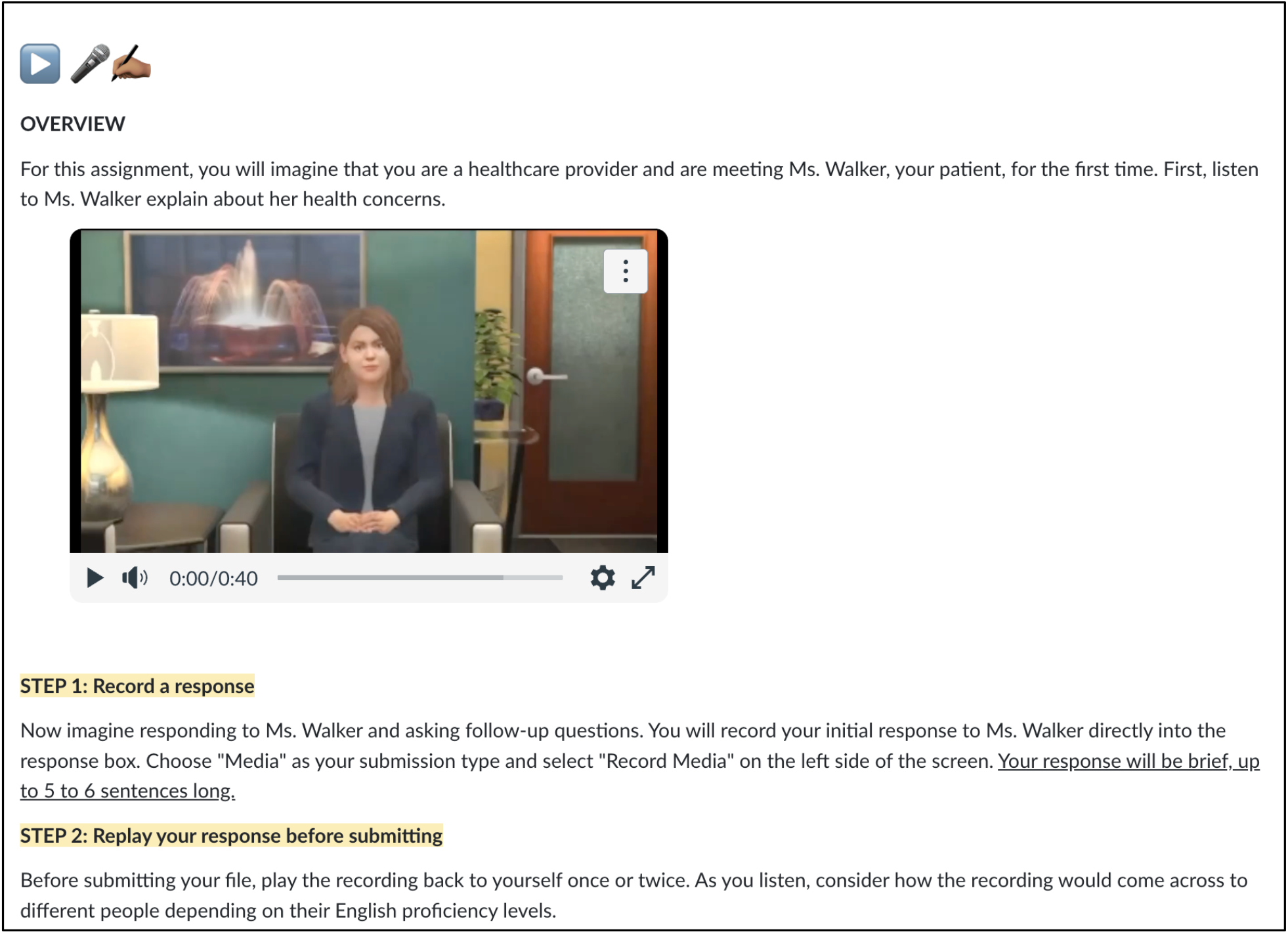
Final Training Activity – Practicing Accommodative Speech Part 1.

**Figure 4.**
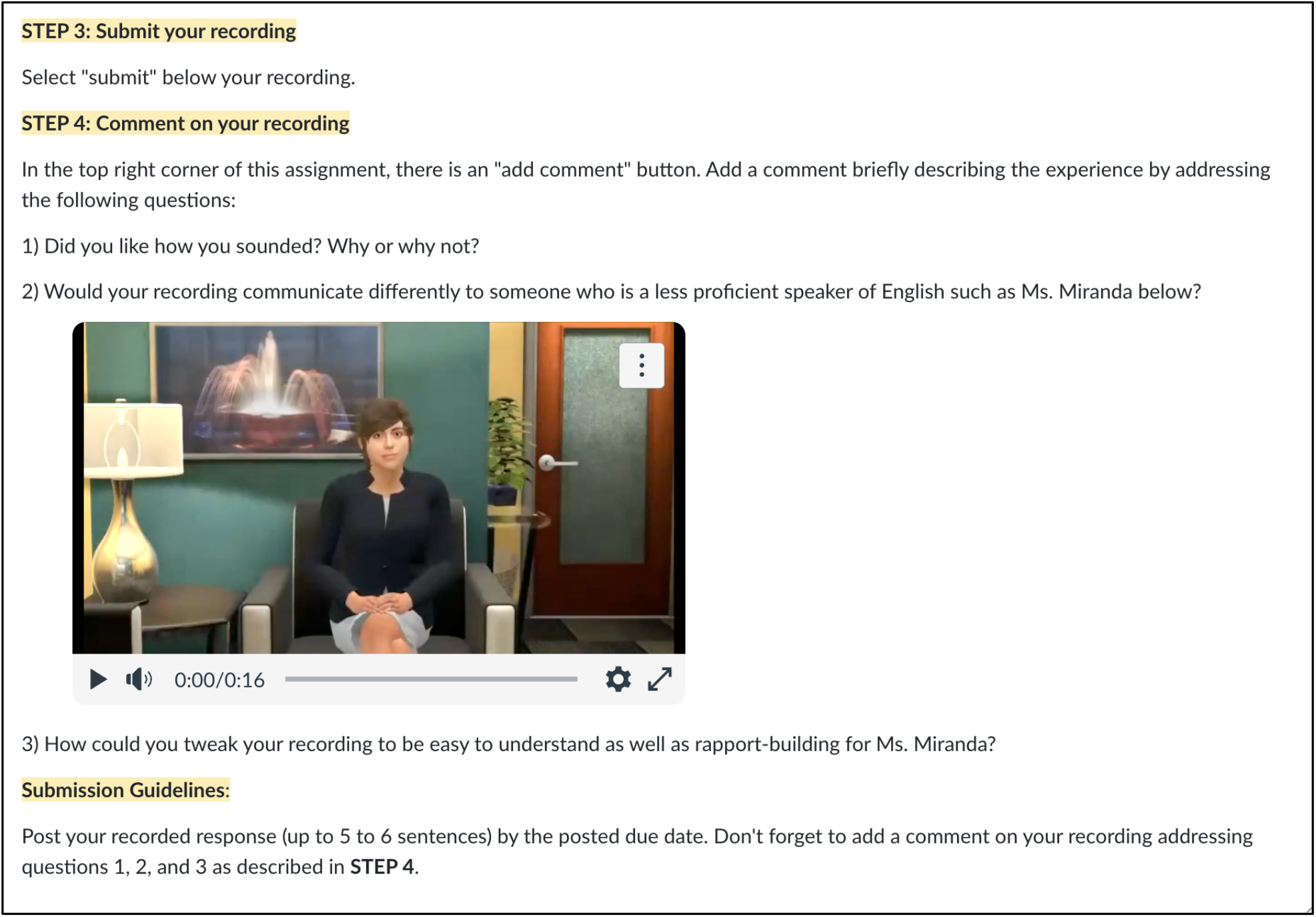
Final Training Activity – Practicing Accommodative Speech Part 2.

#### 2.5.3. Learning Assessment Measures

Learning assessment occurred using the same four multiple-choice knowledge items administered before and after the training. The items addressed (1) characteristics of effective versus ineffective accommodation, (2) the primary purpose of accommodation in interactions involving differences in language proficiency, (3) distinguishing speech accommodation from code-switching or translation, and (4) selecting appropriate accommodation strategies in a clinical scenario (see **Table 2** for details). Following completion of the module only, students answered two questions that rated their 1) confidence in adapting aspects of their speech, including vocabulary, speaking rate, and examples, and 2) their likelihood of using accommodation strategies such as a slower speech rate in future interactions with patients with limited English proficiency. Both items were rated on 6-point scales ranging from 0 (*not at all confident/likely*) to 5 (*completely confident/likely*). The knowledge and confidence items were developed by the authors for this study and were not piloted.

**Table 2.** Percentage of correct knowledge-check responses before and after the module, by site (n = 63).

| Knowledge-check item | Correct answer | Gordon Pre (%) | Gordon Post (%) | ECU Pre (%) | ECU Post (%) |
| --- | --- | --- | --- | --- | --- |
| When clinicians purposefully accommodate speech for a patient with limited language proficiency, which is least characteristic of effective accommodation? | Repeating identical sentences verbatim several times to ensure the message is received | 95.5 | 100.0 | 82.9 | 92.7 |
| Why do clinicians accommodate speech when there is a language proficiency gap? | To increase mutual comprehension while preserving the integrity of core clinical information | 100.0 | 95.5 | 97.6 | 100.0 |
| Which vignette best illustrates speech accommodation rather than code-switching or translation? | A nurse rewords technical terms into simpler English, adds gestures, and pauses to check understanding | 95.5 | 100.0 | 90.2 | 95.1 |
| Which home-care instruction strategy best reflects effective speech accommodation? | Using simplified vocabulary, slower pace, and confirming comprehension through patient feedback | 100.0 | 100.0 | 97.6 | 97.6 |

#### 2.5.4. Instructional Outcome Measures

Our main outcome measure for self-reported changes in accommodation behavior was the self-report Communication Accommodation Behavior Scale (CABS) [26]. Items were rated on 5-point scales ranging from 1 (*strongly disagree*) to 5 (*strongly agree*). We developed CABS as a trait-level instrument that consists of 8 items with two four-item subscales: *facilitative accommodation* which captures prosocial adjustments supporting comprehension (e.g., “I adjust my speech when talking to a less proficient English speaker to facilitate understanding”), and *resistant accommodation*, which captures avoidance or condescension toward lower-proficiency speakers (e.g., “I feel that I sometimes talk down to a less proficient English speaker”). Increased facilitative and decreased resistant scores indicate improved accommodative behavior among clinical students. The CABS was previously validated in a large general adult sample (N= 525). The two subscales showed good internal consistency in validation (α = .87 and .82) [26] and were confirmed as distinct factors via exploratory factor analysis (r = .05 between factors) [26]. In the present sample, internal consistency was acceptable for the facilitative subscale (pre α = .74, post α = .94) and lower for the resistant subscale (pre α = .57, post α = .77). We administered the CABS before and after training. In the post-test, items were reworded to refer to anticipated behavior following the module (e.g., “Now I will adjust my speech…”; “Now I feel that I will sometimes talk down…”). The reworded post-test version has not been separately validated.

### 2.6. Data analysis

All statistical analyses were completed using R version 4.4.3 [28]. Analyses were restricted to participants with matched pre- and post-module surveys and no data were imputed. Among matched participants, there were no missing responses on outcome measures. Pre- and post-training CABS subscale scores were compared using Wilcoxon signed-rank tests. This test was conducted separately for the Facilitative and Resistant subscales within each course (Gordon College and ECU), yielding a test statistic and *p* value for each comparison. Effect sizes were calculated as r = Z/√N. Medians and interquartile ranges (*Mdn, IQR*), rather than means and standard deviations, were reported as the primary descriptive statistics to reflect the non-normal distribution of the data. To test whether pre-post change differed between institutions, we fit a linear mixed-effects model for each subscale, with time (pre, post), course (Gordon College, ECU), and their interaction as fixed effects and a random intercept for participant, using the lme4 and lmerTest packages [29,30]. F tests used Satterthwaite’s approximation for degrees of freedom. Cronbach’s α was computed separately at pre- and post-test using the psych package [31]. Knowledge-check performance was summarized descriptively as percentage correct for each item, by course and time point. Post-training confidence and likelihood-of-use items, collected only at post-test, were summarized descriptively using means and standard deviations (*M, SD*).

## 3. Results

### 3.1. Feasibility

Of 69 students offered the module, 68 (99%) chose it over the alternative assignment (Gordon College: 24 of 24; ECU: 44 of 45). Of these, 64 (94%) completed both the pre- and post-assessment (Gordon College: 22 of 24; ECU: 42 of 44). However, one ECU participant completed both assessments in the incorrect order and was excluded from analysis, resulting in a final analytic sample of 63 (Gordon College: n = 22; ECU: n = 41). Module activity was concentrated in the week after release. Approximated time on the pre- and post-assessments was 14 and 4 minutes, respectively; time spent on the content activities was not recorded.

### 3.2. CABS Subscale Scores

Figure 5 illustrates the changes in CABS scores for both institutions. On the facilitative subscale, scores increased from pre- to post-training in both courses (Gordon College: *Mdn* = 4.25, IQR = 0.75 at pre; *Mdn* = 5.00, IQR = 0.19 at post; ECU: *Mdn* = 4.00, IQR = 0.50 at pre; *Mdn* = 5.00, IQR = 1.00 at post). This increase was statistically significant at both Gordon College (*V* = 12.0, *p* = .002, *r* = .71) and ECU (*V* = 91.5, *p* = .001, *r* = .52). A linear mixed-effects model testing whether this increase differed by course found no significant Time × Course interaction, *F*(1, 61) = 2.21, *p* = .14, indicating that the magnitude of improvement did not differ significantly between institutions.

**Figure 5.**
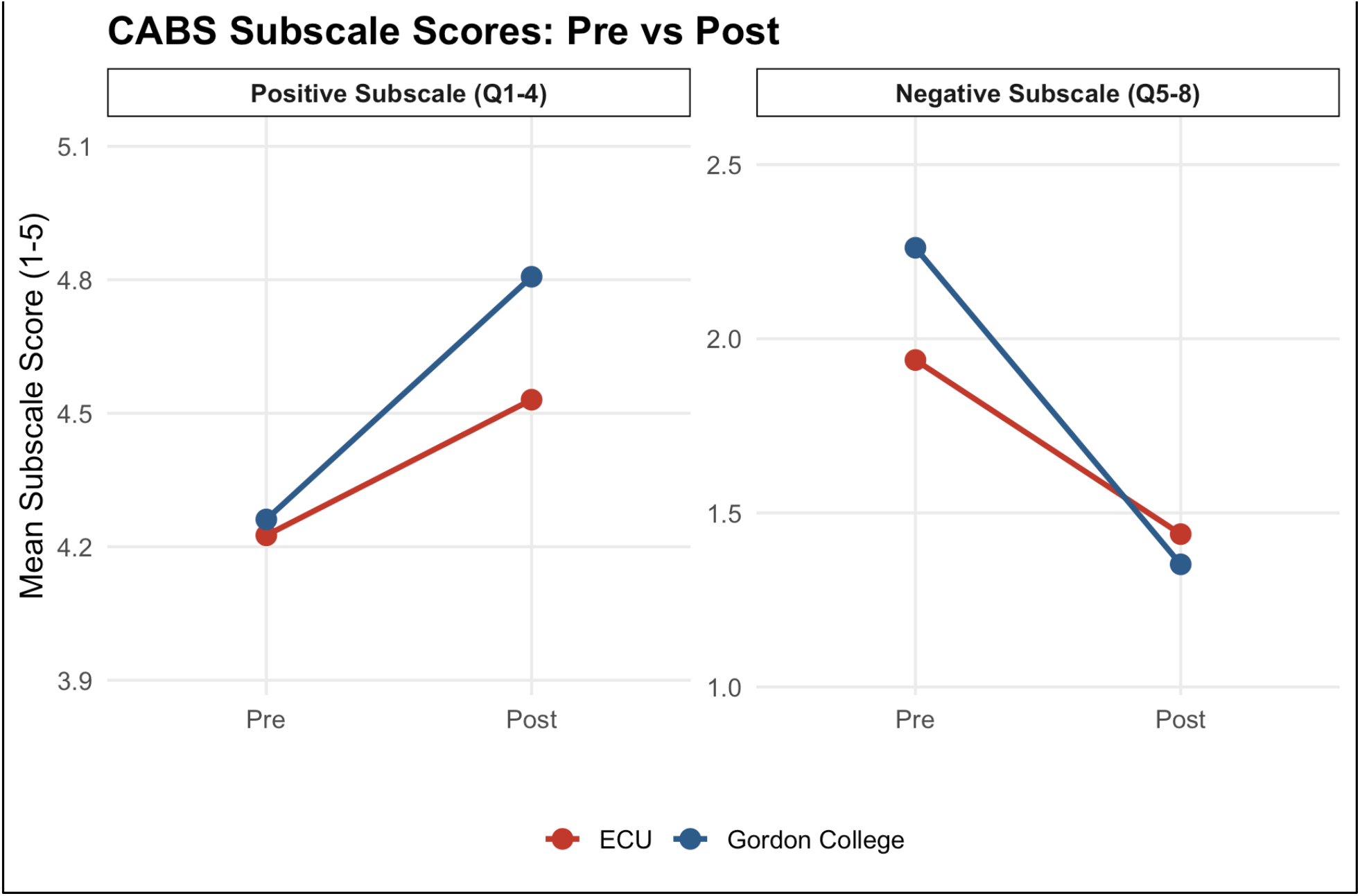
Mean pre- and post-training CABS subscale scores by site (n = 63).

On the resistant subscale, scores decreased from pre- to post-training in both courses (Gordon College: *Mdn* = 2.25, IQR = 0.75 at pre; *Mdn* = 1.25, IQR = 0.44 at post; ECU: *Mdn* = 1.75, IQR = 0.75 at pre; *Mdn* = 1.25, IQR = 0.75 at post). This decrease was statistically significant at both Gordon College (*V* = 224.0, *p* < .001, *r* = .82) and ECU (*V* = 543.0, *p* < .001, *r* = .69). The corresponding mixed-effects model found a significant Time × Course interaction, *F*(1, 61) = 5.35, *p* = .024, indicating a larger reduction in resistance at Gordon College (*M* = 2.26 to 1.35) than at ECU (*M* = 1.94 to 1.44).

### 3.3. Knowledge Check

Percentage correct was high for all four knowledge-check questions in both courses (82.9%– 100% at pre-test and 92.7%–100% at post-test). This pattern suggests a possible ceiling effect and indicates that students already had high content knowledge before completing the module.

### 3.4. Confidence and Likelihood of Future Use

Following training, students reported moderate-to-high confidence in their ability to adapt their speech for a less proficient speaker (Gordon College: *M* = 3.77, *SD* = 0.87; ECU: *M* = 3.27, SD = 0.67, on a 0–5 scale) and high likelihood of using a learned strategy in a future encounter (Gordon College: *M* = 4.32, *SD* = 0.65; ECU: *M* = 4.32, *SD* = 0.72).

## 4. Discussion and Conclusion

### 4.1. Discussion

This study evaluated the development and feasibility of a relatively brief, online communication accommodation training module for speech-language pathology and psychology students, tested at two academic sites on the U.S. East Coast. Results suggest that the module was feasible to deliver in both an undergraduate and a graduate speech-language pathology course, and students showed statistically significant pre-to-post improvement in self-reported communication accommodation. Knowledge scores were high before students engaged in the module and remained high after completion. Overall, the findings provide preliminary support for the module’s feasibility and potential value in health professions education.

Communication Accommodation Theory (CAT) provides a useful framework for interpreting the CABS self-report findings. In interactions with people with a preference for a non-dominant language, uncertainty about communication needs may contribute to inappropriate accommodation, including excessive adjustment, condescension, or avoidance [32]. These responses are reflected in the CABS resistant subscale through items related to “talking down,” using an arrogant tone, or avoiding interaction. CAT also suggests that such responses may occur when speakers rely on broad social categories, such as viewing someone primarily as a LX speaker rather than adapting to the individual’s communication needs [33]. Because facilitative and resistant accommodation are measured as separate dimensions, the changes observed here suggest that the module did more than increase students’ general tendency to adjust their communication. Students reported *greater* use of supportive accommodation while also reporting *less* condescension and avoidance. From a CAT perspective, this pattern may reflect a shift toward more appropriately calibrated and individualized communication. This distinction is important because effective accommodation involves not only adjusting communication *more* but adjusting it in ways that are responsive to the individual listener. The module’s behavioral rehearsal activity, in which students recorded and evaluated their own speech, may have contributed to these changes by requiring application rather than passive exposure; in future studies, these recordings could be coded for observable accommodation strategies. Although both sites improved, the decrease in resistant accommodation was larger at Gordon College than at ECU. Gordon College students began with higher resistant scores and therefore had more room for change. The sites also differed in academic level, program, and prior clinical exposure, so the present design cannot determine which of these factors accounts for the difference.

There are several implications for the design and evaluation of communication training when considered through the Kirkpatrick framework [23]. At Level 1, students reported moderate-to-high confidence in their ability to adapt their speech and a high likelihood of using accommodation strategies in future interactions. Although these measures do not represent a traditional satisfaction-based reaction assessment, they suggest that learners perceived the strategies as relevant and potentially usable in future clinical contexts. For a relatively brief online intervention, this perceived readiness for future use is meaningful because learners could envision applying what they encountered in the module beyond the immediate learning experience. Similar health professions e-learning research has used the Kirkpatrick framework to examine learner perceptions and confidence alongside subsequent learning and behavioral outcomes, demonstrating the value of considering these outcomes as related but distinct indicators of training effectiveness [34].

At Level 2, the findings were more complex. Knowledge-check performance was already high before training and remained high following completion, indicating, as previously stated, a likely ceiling effect. The knowledge items primarily assessed recognition of foundational accommodation concepts, whereas the module itself required learners to observe communication, discriminate between accommodative and nonaccommodative examples, apply strategies in their own speech, and reflect on their performance. This distinction is important from an instructional design perspective. Merrill [35] proposes that learning is promoted when learners activate existing knowledge and then engage in demonstration, application, and integration. Students in the present study appeared to enter the module with substantial foundational knowledge, suggesting that the instructional value of the experience may have been less about acquiring additional information and more about applying and reconsidering that knowledge in context. Future Level 2 assessments should therefore closely align with these intended learning outcomes by requiring learners to evaluate ambiguous clinical scenarios, distinguish underaccommodation from overaccommodation, select strategies appropriate to an individual patient, or explain what additional information they would need before making a communication adjustment. Rather than simply adding additional instructional content, future development should emphasize increasingly complex opportunities for clinical judgment, individualized decision-making, behavioral rehearsal, and assessment of observable performance.

The Level 3 findings are encouraging because facilitative accommodation increased and resistant accommodation decreased significantly at both sites following the module, suggesting a shift in learners’ self-reported behavioral orientation toward communication accommodation. Because post-test CABS items were worded prospectively, these scores reflect anticipated rather than current accommodation behavior. However, because the CABS is a self-report measure, these findings do not establish observable behavior change in clinical practice (Kirkpatrick Level 3 – Behavior). This distinction is consistent with recent application of the Kirkpatrick framework in allied health education, where behavioral outcomes were examined through reported application and transfer following training and were assessed longitudinally beyond the immediate learning experience [34]. Future evaluation of the present module should similarly extend Level 3 to directly measure performance through simulation, standardized patient encounters, mixed-reality environments, or authentic clinical interactions. For example, mixed-reality tools such as Mursion could be integrated in future iterations of the module [15].

The recording and reflection activity already incorporated into the module provides a foundation for this progression because learners practiced accommodative speech, listened to their own communication, and considered how it might be perceived by listeners with different levels of proficiency. Such opportunities for application and reflection are consistent with instructional approaches that move learners beyond exposure to information toward active use and integration of knowledge [35].

Finally, the Kirkpatrick framework [23] highlights an important distinction between knowing recommended communication strategies and applying them appropriately in context. Students demonstrated substantial baseline knowledge, yet their self-reported accommodation still shifted, suggesting that communication accommodation training should focus on helping students recognize their own tendencies and calibrate adjustments to individual listeners. Level 4 outcomes were beyond the scope of this feasibility study; future research could examine whether changes in accommodation contribute to patient understanding, rapport, or trust.

### 4.2. Limitations

Our study has several limitations. First, the CABS is a self-report measure, so the scores may be influenced by a so-called social desirability bias [36]. It is also unclear if the scores relate to actual changes in clinical communication. Thus, longitudinal studies would be helpful as well as ratings of students’ communication by people with a language preference other than English. Additionally, our sample was predominantly female, White, and first-language English speakers in speech-language pathology and psychology programs. Future studies should examine feasibility in more diverse samples and in other health professions, such as nursing, medicine, and physician assistant programs. The knowledge questions may have been too easy given the high pre-training scores, so revised items should be more difficult to better capture learning gains. Internal consistency of the resistant subscale was low at pre-test, so these scores should be interpreted with caution. Also, without a control group, we cannot rule out testing effects or other influences unrelated to the module. Finally, the module focused on capability rather than motivation. Future work could address motivation to change communication styles as well as structural or policy approaches to promote accommodation.

### 4.3. Conclusion

We presented outcomes of a brief online communication accommodation module that was associated with short-term improvements in self-reported communication accommodation. Knowledge scores were high before and after training, suggesting the module may be more valuable for application than knowledge acquisition. Students also reported moderate-to-high confidence and likelihood of future use. Together, these findings offer preliminary support for a scalable, low-cost training approach that health professions programs could adopt to better prepare students for patients with limited English proficiency.

### 4.4. Practice implications

Health professions programs can embed a brief online module with self-recorded practice into existing courses to help students calibrate their communication with LX patients. Assessments should target clinical judgment rather than recognition of concepts.

## Supporting information

SupplementaryFile1_CRe-DEPTH_Reporting_Checklist

## Data Availability

De-identified data supporting the findings of this study are available from the corresponding author upon reasonable request, subject to applicable institutional and ethical requirements.

## CRediT authorship contribution statement

**Kathrin Rothermich:** Conceptualization, Methodology, Investigation, Data curation, Formal analysis, Funding acquisition, Project administration, Supervision, Writing – original draft, Writing – review & editing.

**Rebecca Clark-Stallkamp:** Formal analysis, Visualization, Writing – original draft, Writing – review & editing.

**Rose P. Baker-Iyore:** Methodology, Resources, Writing – review & editing.

**Charlene Cuatico:** Conceptualization, Methodology, Resources.

**Joseph G. L. Lee:** Conceptualization, Funding acquisition, Writing – review & editing.

**Susan C. Bobb:** Conceptualization, Methodology, Investigation, Funding acquisition, Project administration, Writing – review & editing.

## Declaration of generative AI and AI-assisted technologies in the manuscript preparation process

The authors used ChatGPT (OpenAI) to assist in the creation of Figure 1, a conceptual diagram illustrating the integration of the Kirkpatrick Evaluation Model into an ADDIE-based instructional design framework. The figure was based on author-developed content and was reviewed, edited, and verified by the authors prior to submission. The authors take full responsibility for the content of the figure and the manuscript. In addition, NotebookLM (Google) was used to generate an educational podcast incorporated into the training module evaluated in this study. The podcast served as instructional content within the intervention and was not used in the preparation, analysis, or writing of the manuscript.

