## SupplementaryFile1_CRe-DEPTH_Reporting_Checklist for "Development and Feasibility of a Communication Accommodation Training Module for Health Profession Students"

**Supplementary File 1: CRe-DEPTH Reporting Checklist**

**Manuscript:** Development and Feasibility of a Communication Accommodation Training Module for Health Profession Students

This checklist shows where the manuscript reports each of the 12 items in the Criteria for Reporting on Development and Evaluation of Professional Training interventions in Healthcare (CRe-DEPTH; Van Hecke A, Duprez V, Pype P, Beeckman D, Verhaeghe S. Nurse Educ Today 2020;84:104254. https://doi.org/10.1016/j.nedt.2019.104254).

Page numbers refer to the manuscript as submitted.

**Checklist**

| No. | CRe-DEPTH item | Reported on page / section |
|---|---|---|
| | Development of the training | |
| 1 | Aim or objectives of the training | p. 5 (Module description); p. 6 (Structure and procedure) |
| 2 | Underlying theoretical framework | pp. 3–4 (Introduction); p. 5 (Module description) |
| 3 | Developmental process | p. 5 (Intervention development); p. 6 (Figure 1) |
| 4 | Target population and setting | p. 4 (Participants); pp. 4–5 (Table 1) |
| 5 | Educational resources | p. 5 (Module description); pp. 7–8 (Content). Development costs not reported |
| | Characteristics of the training | |
| 6 | Content of the training | pp. 7–9 (Content; Figures 2–4) |
| 7 | Format | p. 5 (Module description); p. 10 (Results: Feasibility) |
| 8 | Didactic methods | pp. 7–8 (Content) |
| 9 | Tailoring of the training | p. 5 (Module description). Not tailored; content identical across sites and academic levels |
| | Characteristics of the providers | |
| 10 | Providers of the training | p. 5 (Module description). Self-directed online module; course instructors introduced it and posted reminders but gave no feedback |
| | Assessment of the training outcomes | |
| 11 | Measured outcomes | pp. 6–7 (Module structure); pp. 9–10 (Measures); pp. 10–12 (Results) |
| 12 | Assessment method, including validity and reliability | pp. 9–10 (Learning assessment measures; Instructional outcome measures); p. 10 (Data analysis) |
